# STRIVE-ON: Safety and ToleRability of GTX-104 (Nimodipine Injection for IV Infusion) ComparEd with Oral Nimodipine in Patients Hospitalized for Aneurysmal Subarachnoid Hemorrhage (aSAH): a Prospective, Randomized Trial (STRIVE-ON)

**DOI:** 10.1101/2024.09.25.24314408

**Authors:** A Choi, SH-Y Chou, AF Ducruet, WT Kimberly, RL Macdonald, AA Rabinstein

## Abstract

Oral nimodipine is the only drug approved in North America for treatment of patients with aneurysmal subarachnoid hemorrhage (aSAH). However, bioavailability is variable and frequently poor, leading to fluctuations in peak plasma concentrations that cause dose-limiting hypotension. Furthermore, administration is problematic in patients who cannot swallow capsules. An oral liquid formulation exists but causes gastrointestinal complications. An intravenous nimodipine formulation (GTX-104) has been developed that has bioavailability approaching 100% and is not affected by feeding or gastointestinal absorption. GTX-104 causes less hypotension and has more consistent peak plasma concentrations than oral nimodipine in healthy human volunteers.

Herein we describe the protocol of a prospective, randomized, open-label safety and tolerability study of GTX-104 compared to oral nimodipine in patients with aSAH (STRIVE-ON, NCT05995405). Inclusion and exclusion criteria match the prescribing information for oral nimodipine and include adult patients with aSAH of all Hunt and Hess grades who can receive investigational product within 96 hours of aSAH. Subjects at imminent risk of death are excluded. Subjects are randomized 1:1 to GTX-104 or oral nimodipine for up to 21 days. The primary endpoint is the proportion of subjects in each group with clinically significant hypotension, defined as hypotension requiring any medical treatment, with a reasonable likelihood of being due to investigational product as determined by an independent, blinded endpoint adjudication committee. No statistical analysis of the endpoint is planned. Secondary endpoints include all episodes of hypotension, all adverse events, delayed cerebral ischemia, rescue therapy and suicidal ideation. Clinical and health economic outcomes include quality of life using the EQ-5D-3L, modified Rankin scale at 30 and 90 days after aSAH and hospital resource use. The planned sample size is 100 subjects across 25 sites in the United States and Canada.

**DETAILS PAGE:**

1. We confirm that manuscript complies with all instructions to authors.
2. We confirm that authorship requirements have been met and the final manuscript is approved by all authors.
3. We confirm that this manuscript has not been published elsewhere and is not under consideration by another journal. It is planned to post it on
4. We confirm adherence to ethical guidelines and indicate ethical approvals and use of informed consent.
5. All conflicts of Interest for all authors are disclosed.
6. We confirm the use of the Standard Protocol Items: Recommended for Interventional Trials (SPIRIT) checklist.
7. The source of funding for the study is disclosed. It is Acasti Pharma.

## INTRODUCTION

This report adheres to the Standard Protocol Items: Recommended for Interventional Trials (SPIRIT).^1^ Nimodipine is the only drug approved for administration to patients with aneurysmal subarachnoid hemorrhage (aSAH).^2^ It is a dihydropyridine molecule that antagonizes L-type calcium channels that are located predominately on vascular smooth muscle. Two oral formulations (capsules, liquid solution) are available in the United States (US) whereas tablets and an intravenous (IV) formulation are available in the European Union and some other countries.

There are problems with the oral nimodipine capsules, including poor and variable bioavailability, high first-pass effect, risk of inadvertent IV injection and need to aspirate the liquid from the capsules to administer to patients who cannot swallow if the oral liquid solution is not available.^3,4^ The oral nimodipine solution causes diarrhea.^5^ These lead to poor compliance and unpredictable episodes of hypotension.^6–10^ Hypotension remains the only important and frequent adverse effect of nimodipine.^11^ The nimodipine prescribing information reports 5% of patients developed hypotension at the approved dosage. Another clinical trial reported oral nimodipine caused hypotension (defined as decrease in systolic blood pressure [BP] >20 mm Hg, diastolic BP >10 mm Hg, or systolic BP ≤90 mm Hg, confirmed by 3 consecutive readings and requiring medical treatment) in 10% of patients.^12^ The frequency of dose reductions and discontinuations of oral nimodipine due to hypotension is even higher in observational studies and is up to 44%.^7,9,10,13^

Although IV nimodipine might avoid some of these problems, there are limitations of the IV formulation of nimodipine. The safety and efficacy of nimodipine is based on the oral formulations. The IV solution was not studied as much in the nimodipine clinical trials and metaanalysis of nimodipine wrote that there was insufficient data to define the benefits of IV nimodipine.^14^ In addition to limited sample size, the IV nimodipine dose regimen is a continuous infusion and does not produce the saw-tooth pharmacokinetic profile of oral nimodipine. It is not know if the pharmacokinetic profile is important for the efficacy of nimodipine. Additionally, each 50 mL bottle of the IV formulation available in the European Union contains 10 g ethanol (96%) and 10 mg nimodipine. At the prescribed dosage this is almost a bottle of wine per day. Hence, liver toxicity is a concern. This formulation has to be given through a central line because it causes vein irritation, pain and inflammation.^15^ Finally, hypotension is reported in about 30% of patients.^6,16–20^

A new formulation of IV nimodipine, GTX-104, could overcome the limitations of existing nimodipine formulations. Putative advantages of GTX-104 include less inter-and intrapatient variability in pharmacokinetics (PK) at least in healthy human volunteers and thus less hypotension, almost 100% bioavailability because of no food effect or first pass metabolism and better compliance due to no difficulty administering to patients who cannot swallow.

## PHASE 1 CLINICAL DATA

GTX-104 has been administered to 104 healthy human volunteers in 2 studies (unpublished data, Acasti Pharma). A phase 1, single center, randomized, 2-period cross over PK study assessed GTX-104 and oral nimodipine capsules, which are the reference standard, in 58 subjects.

GTX-104 was administered for 72 hours as a continuous infusion of 0.15 mg/hour with a 30 minute bolus infusion of 4 mg every 4 hours. Nimodipine capsules were administered orally at a dose of 60 mg (2 30 mg capsules) for 72 hours. The 2 products demonstrated similar results for the 2 primary endpoints (maximum blood concentration after the first dose and the area under the concentration-time curve on the 3^rd^ day), as measured by the ratio and 90% confidence interval (CI) of their geometric means: 92% (90% CI: 82 - 104%) and 106% (90% CI: 99 - 114%), respectively. The secondary PK parameters (daily maximum concentration at steady-state and time to maximum concentration) were also the same for the 2 formulations. The variability in all PK parameters was less for GTX-104 compared to oral nimodipine. The average oral bioavailability for nimodipine capsules was 7%.

## RATIONALE

GTX-104 approval will be sought through the 505(b)(2) regulatory pathway and will rely on the previous findings of safety and effectiveness for the reference drug, oral nimodipine. Thus, a single safety study for registration is planned since there is extensive experience with nimodipine over the last 4 years in thousands of patients where the only important safety issue is hypotension.21,22The reasons that demonstration of efficacy of GTX-104 would not be necessary include that effectively the same dose as the efficacious oral formulation is being administered only by a different route of administration. In addition, oral nimodipine is standard of care so an efficacy study would probably require a non-inferiority design and in the current medical care system, this will never be done.

Given the 505(b)(2) pathway, this protocol mimics the original pivotal oral nimodipine phase 3 clinical trials. Thus, the inclusion and exclusion criteria reflect a combination of considerations related to the oral nimodipine prescribing information and to general clinical trial design. We enroll the full spectrum of aSAH severity based on the Hunt and Hess scale because this scale was used in the original clinical trials, even though the clinical trials that were the basis for approval varied with regard to grades included and doses used.^23–26^

## METHODS

### General Design

This safety study is a prospective, open-label, randomized (1:1 ratio), parallel group study of GTX-104 compared with oral nimodipine, in subjects with aSAH. Randomization is by central interactive response technology. There is no blinding. Approximately 100 subjects will be enrolled (approximately 50 subjects in each treatment group) at approximately 25 sites in the US and Canada.

### Objectives and Endpoints

The objectives are to evaluate the safety and tolerability and clinical and health economic outcomes of patients with aSAH treated with GTX-104 compared to those treated with oral nimodipine.

The primary endpoint is the incidence (% or proportion) of subjects with at least 1 episode of clinically significant hypotension with a reasonable possibility that GTX-104/oral nimodipine caused the event, according to the blinded Endpoint Adjudication Committee (EAC). Secondary safety endpoints include the duration and total number of episodes of clinically significant hypotension (investigator opinion), the incidence and severity of adverse events, delayed cerebral ischemia, rescue therapy and suicidal ideation. Clinical and health economic outcomes include hospital and intensive care unit lengths of stay, quality of life and modified Rankin scale (mRS).

### Study Population

This reflects the pivotal oral nimodipine studies and the current indication for nimodipine.^2,14^ Oral nimodipine originally was approved for Hunt and Hess grades 1 to 3.^27^ This was expanded to all Hunt and Hess grades in 1996. This study therefore enrolls aSAH patients with any Hunt and Hess grade at randomization. Although the primary clinical grading uses the Hunt and Hess scale, both the Hunt and Hess and World Federation of Neurological Surgeons (WFNS) grading scales are considered by the American Heart Association/American Stroke Association, to be simple, validated scales for the initial assessment of the clinical severity of SAH.^2,27,28^ The WFNS scale is widely used in the United States and has better inter and intraobserver reliability than the Hunt and Hess scale so it also will be assessed before randomization.^29^

Study subjects will be male or female patients ≥18 years of age with a diagnosis of aSAH based on computed tomography and angiography (computed tomographic, magnetic resonance or digital subtraction angiography). They must have a Hunt and Hess score from 1 to 5 just prior to randomization. Consent will be obtained. The investigational product (IP) must be started within 96 hours from the onset of the aSAH, as suggested in the prescribing information.

The exclusion criterion which is directly from the nimodipine prescribing information, is patients receiving strong inhibitors of CYP3A4.^21,22^

Also excluded are patients with a history of recurrent syncope or hypotension, those requiring cardiopulmonary resuscitation within 4 days prior to randomization and patients with second-or third-degree atrio-ventricular block or bradycardia (heart rate ≤50 bpm) prior to randomization. These are mentioned as warnings in the prescribing information and are exclusions since these may interfere with the safety assessments. Regulatory advice was to exclude patients with severely impaired liver function since they have impaired first pass effect and decreased liver metabolism (cirrhosis [Child-Pugh class B and C] or alanine aminotransferase and/or aspartate aminotransferase more than 2.5 times the upper limit of normal). These patients can have high and unpredictable blood nimodipine concentrations leading to hypotension. Patients with a history of malabsorption syndrome, recent ileus (in the last 3 months) or other gastrointestinal conditions that would interfere with absorption of nimodipine, in the opinion of the investigator, are not eligible.

The protocol limits patients to 12 doses (or a total of 720 mg) of oral nimodipine (as a solution or capsules) as part of the standard of care for the ruptured aneurysm prior to randomization. Earlier versions of the protocol limited pre-study nimodipine to 5 doses (300 mg) because of concerns that longer times could limit the duration of exposure to GTX-104. This turned out not to be a problem so the number of doses allowed was increased. Exclusion criteria based on general clinical trial principles include patients who are at imminent risk of death and/or have do not resuscitate orders and patients with a severe or unstable concomitant condition or disease other than what may be attributed to the aSAH that, in the opinion of the investigator, may increase the risk associated with study participation or nimodipine administration, or may interfere with the interpretation of study results.

### Study Procedures

The study has pre-randomization (screening), treatment and follow-up periods (**Table 1**, **Figure**). Sites were selected based on many factors including number of aSAH cases per year, neurocritical care and neurovascular surgery infrastructure and experience, clinical trials infrastructure and experience, able and willing to comply with the study protocol, budget and overall responsiveness or interest in the study. The study was approved by each site’s local or centralized ethics board. Regulatory and ethical aspects of the study adhere to consensus ethical principles derived from international guidelines including the Declaration of Helsinki and Council for International Organizations of Medical Sciences International Ethical Guidelines and applicable International Conference on Harmonization Good Clinical Practice Guidelines.

**Figure:**
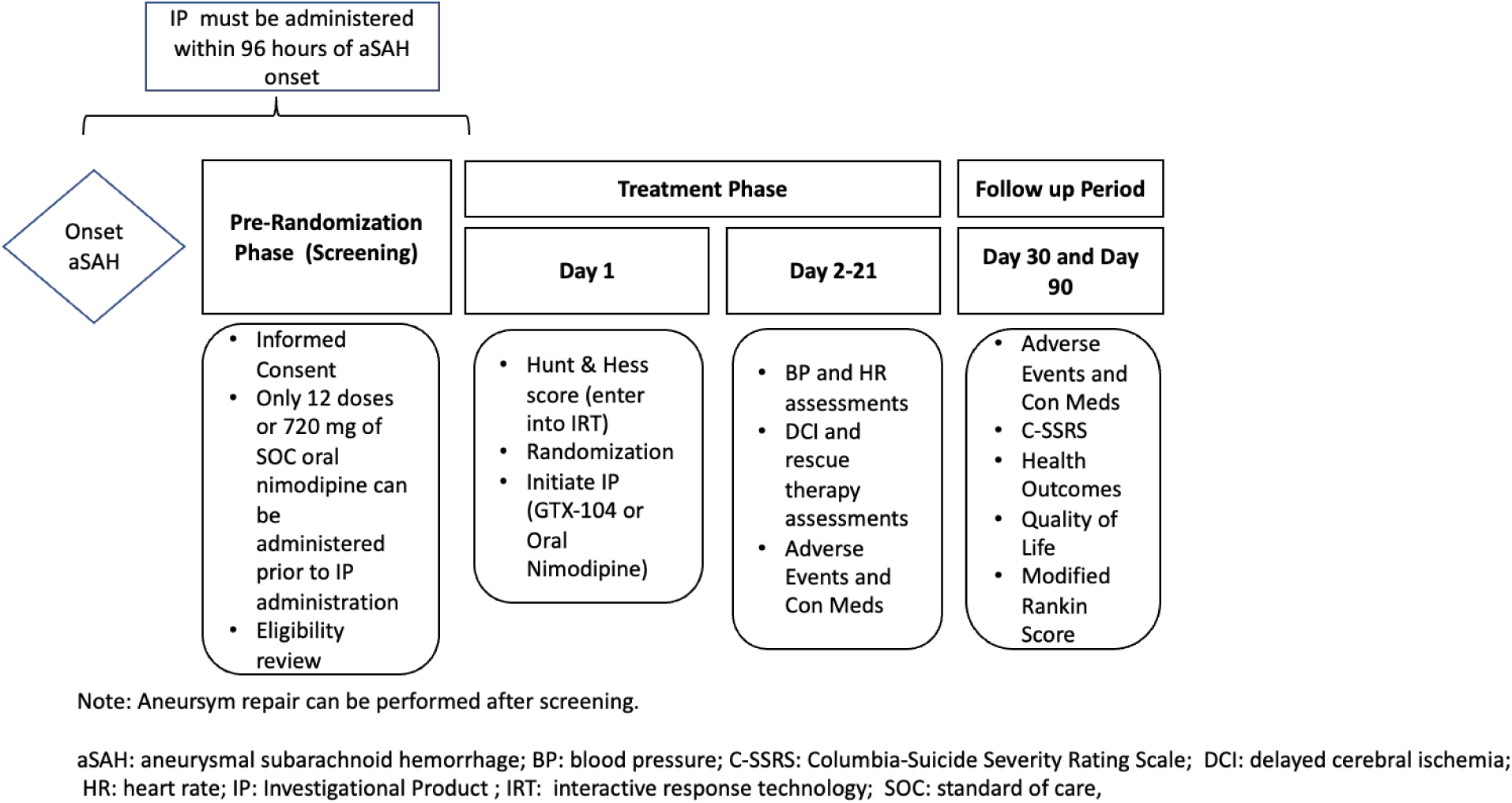
Flow Chart of Study Protocol Basics

**Table 1:**
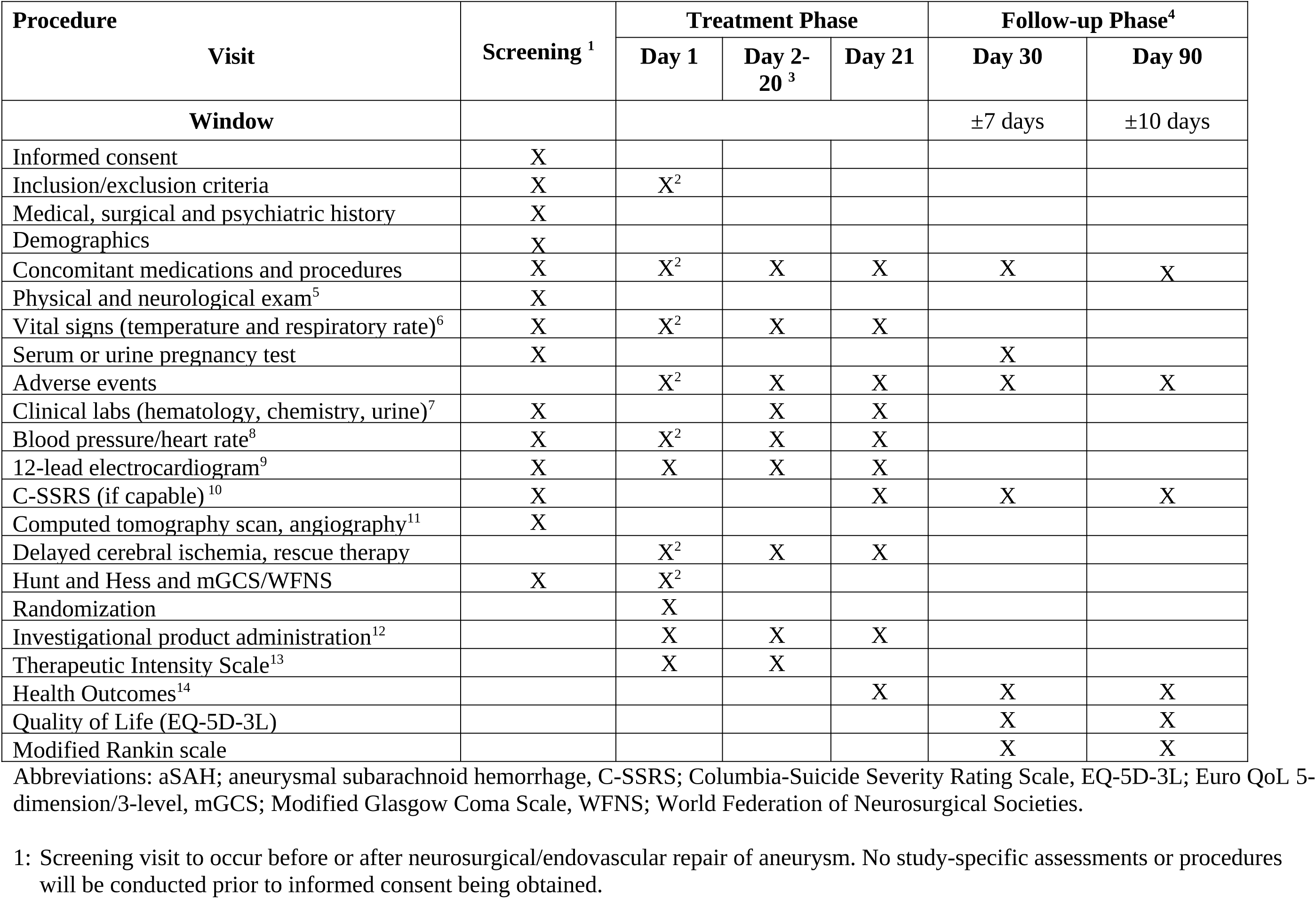
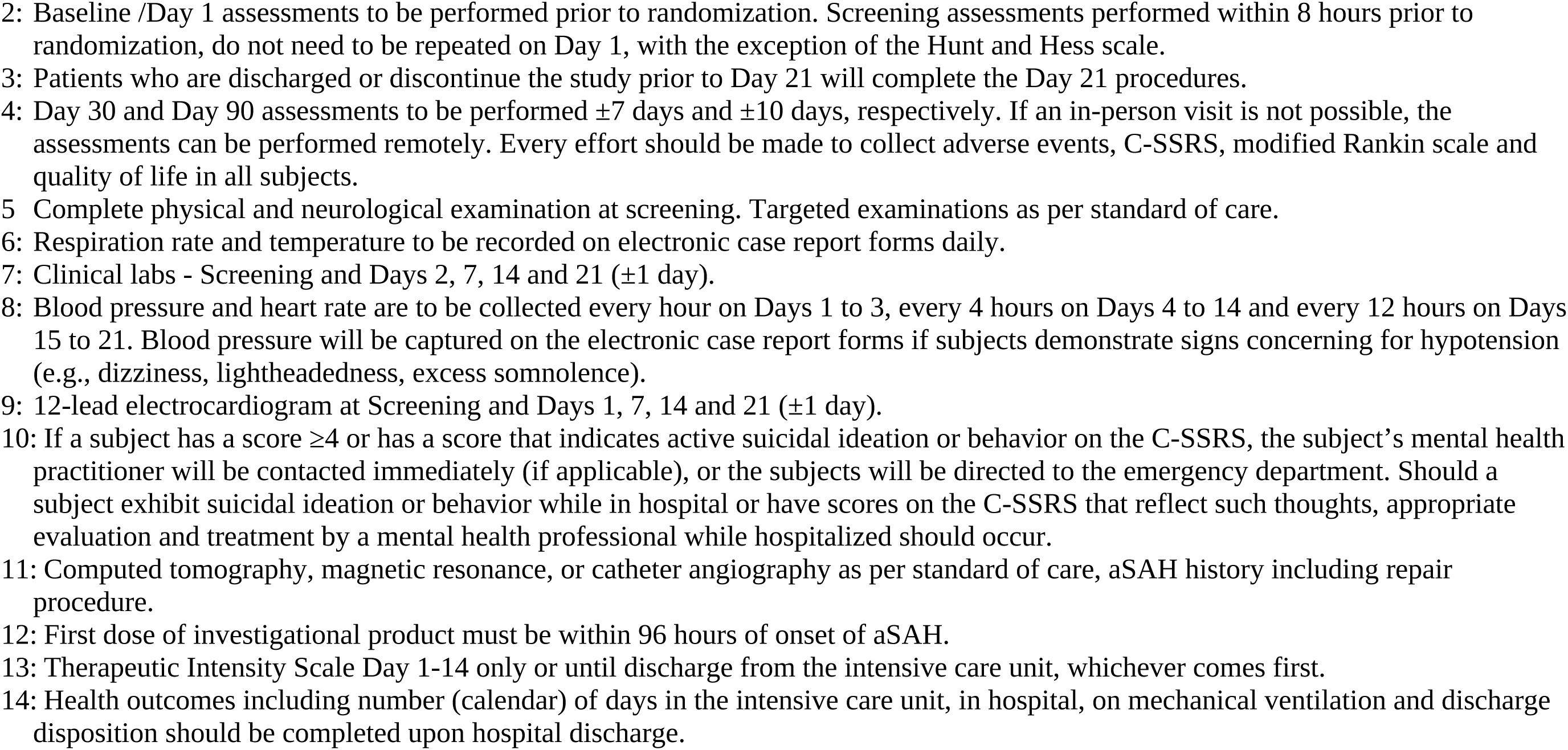
Schedule of Activities.

The study protocol does not require that the ruptured aneurysm be repaired. If the aneurysm is treated, which occurs in the vast majority of the patients who are eligible for this study, neurosurgical clipping or endovascular aneurysm occlusion are acceptable. The timing of aneurysm repair also is not specified for this study. Randomization will be in a 1:1 ratio to receive either GTX-104 or oral nimodipine for up to 21 days. Randomization will be stratified by Hunt and Hess score (1-3 versus 4-5) and age (≤ 59 versus

> 59 years). Additional data collected pre-randomization is what usually is documented in aSAH patients (**Table 1**). The Columbia-Suicide Severity Rating Scale (C-SSRS) will be assessed in all subjects who are able to provide the information. The C-SSRS will also be obtained at the end of the treatment period and at the Day 30 and 90 follow-up visits. The C-SSRS is being assessed because the Federal Drug Administration (FDA) issued a draft guidance in 2012 stating that "prospective suicidal ideation and behavior assessments should be carried out in all clinical trials involving any drug being developed for any psychiatric indication, as well as for all antiepileptic drugs and other neurologic drugs with central nervous system activity…".^30^

The treatment period begins at randomization (Day 1) and continues up to Day 21 or until the subject is transferred to a location that cannot administer IP (e.g., discharge, regular hospital ward), whichever comes first. The focus of this period is documentation of adverse events and hypotension. Heart rate and BP will be measured every hour from Day 1 to Day 3, every 4 hours from Day 4 to Day 14, and every 12 hours from Day 15 to Day 21. In addition, BP will be captured if subjects demonstrate symptoms or signs or radiologic or laboratory findings concerning for hypotension (e.g., dizziness, lightheadedness, excess somnolence, electrocardiogram changes).

Follow-up continues until the Day 90 assessments are completed. There also is a study visit at Day 30. Specific outcomes measured in this period are the modified Rankin scale, quality of life (EQ-5D-3L) and health economic outcomes (days in the intensive care unit, in hospital and on mechanical ventilation and discharge disposition).^31,32^

### Investigational Products

One arm of the study is administered GTX-104 which is a sterile solution containing nimodipine (2 mg/mL). The nimodipine is solubilized in polysorbate 80 micelles, 1.26% weight/volume of ultrapure alcohol and sterile water. It is diluted in normal saline prior to administration. Polysorbate 80 is an excipient already used in other IV drugs such as amiodarone and some chemotherapy drugs including docetaxel, epoetin/darepoetin and fosaprepitant.^33,34^ GTX-104 can be given through a peripheral or central venous catheter. All IV apparatus used cannot contain polyvinyl chloride. Nimodipine binds to polyvinyl chloride and polysorbate 80 can leach diethylhexyl phthalate from polyvinyl chloride.^35^

The other arm of the study receives oral nimodipine which is a gelatin capsule containing nimodipine, 30 mg. Oral nimodipine is preferably administered in a fasting state (i.e., not less than 1 hour before or 2 hours after a meal). This is recommended in the prescribing information but anecdotally is not always adhered to in practice. If a subject cannot swallow the nimodipine capsule, the instructions in the package insert are followed.

Subjects can receive liquid oral nimodipine solution prior to consent and randomization but not afterwards. This is because this study is using a PK bridging strategy to gain FDA approval and the dose of GTX-104 was selected to match the PK of the oral capsules which are the reference standard from a regulatory perspective.

The management of subjects is left to the centers who are to use their standard of care.

### Primary Endpoint -Hypotension and Dose Reduction

The only common side effect of nimodipine is hypotension. Therefore, hypotension is the primary endpoint of the study. Hypotension is defined as a decrease in systolic BP >20 mm Hg, diastolic BP >10 mm Hg or a systolic BP ≤100 mm Hg, confirmed by 2 consecutive readings within 5 minutes.

Two categories of hypotension have been defined:

> Not clinically significant: not requiring any medical treatment (pharmacotherapy or other intervention).

> Clinically significant: requiring medical treatment, including but not limited to IV fluids, postural changes, dose reduction of IP, interruption of antihypertensive medications, prescription of vasopressors, increasing dose of a vasopressor or addition of a new vasopressor.

If the site investigator deems it necessary, administration of IP can be interupted or the dose of IP can be reduced. The dose reduction should follow the recommendations in the FDA label.^21^ The most recent label states that some patients, such as those taking moderate or weak CYP3A4 inhibitors, may require a nimodipine dose reduction in case of hypotension. The only specific statement about dose reduction pertains to those with severely disturbed liver function in which the dosage may be reduced to 30 mg every 4 hours. Furthermore, guidelines for management of aSAH state that the recommended dose should be given “even in the setting of nimodipine-induced hypotension that can be managed with standard medical interventions. However, if nimodipine causes significant BP variability, temporary stoppage may be necessary.”^2^

Some investigators routinely administer oral nimodipine, 30 mg every 2 or 4 hours. Administration every 2 hours clearly does not follow the FDA label. In one of the protocol amendments, we wrote that dosing every 2 hours was acceptable. However, the FDA asked us to justify this, which we found difficult, so this was removed.

A principle of this study is to consider the dose regimens of the 2 drugs as if they are the same and to manage side effects the same way in each study group. For the conversion of doses between GTX-104 and oral nimodipine, 30 mg oral nimodipine is equal to a 2 mg bolus of GTX-104.

### Secondary Endpoints

The first secondary endpoint is self explanatory. It is all episodes of clinically significant hypotension and their duration, as reported by the investigators regardless of suspected cause. This will be presented as number of subjects and number of events per subject with descriptive statistics of the durations.

Reporting of adverse events follows good clinical practice guidelines and uses the incidence and severity of adverse events based on the National Cancer Institute Common Terminology Criteria for Adverse Events (Version 5.0).

### Delayed Cerebral Ischemia and Rescue Therapy

The definition of delayed cerebral ischemia (DCI) is from Vergouwen, et al.^36^ Rescue therapy may be administered in the event of angiographic vasospasm and/or DCI according to the site standard of care. Rescue therapy will be defined as induced hypertension, selective intraarterial infusion of vasodilator drugs or balloon angioplasty. Information about angiographic vasospasm, DCI and rescue therapy will be captured for this study. The protocol states that in general, off-label use of other drugs for the treatment of angiographic vasospasm and DCI, such as intra-arterial nicardipine/verapamil, initiation of statin therapy or infusion of magnesium for the purposes of achieving supratherapeutic blood magnesium concentrations are discouraged and should be avoided. This is consistent with guidelines.^2^ It is recognized that some centers will use these drugs off-label for these indications.

### Concomitant Medications and Interventions

Concomitant administration of IP (either GTX-104 or oral nimodipine) and strong CYP3A4 inhibitors is contraindicated, based on the nimodipine prescribing information. Strong inhibitors of CYP3A4 include some macrolide antibiotics (e.g., clarithromycin, telithromycin), some anti-human immunodeficiency virus protease inhibitors (e.g., delaviridine, indinavir, nelfinavir, ritonavir, saquinavir), some azole antimycotics (e.g., ketoconazole, itraconazole, voriconazole) and some antidepressants (e.g., nefazadone). Grapefruit juice may potentiate the hypotensive effect of nimodipine. The BP lowering effect may last for 4 or more days after an intake of grapefruit juice so drinking or eating grapefruits/grapefruit juice is not recommended while taking nimodipine.

Strong CYP3A4 inducers (e.g., rifampin, phenobarbital, phenytoin, carbamazepine, St John’s wort) should be avoided in patients prescribed nimodipine since they can lower the plasma concentration of nimodipine.

Selected concomitant medications and procedures will be recorded. These include medications with cardiovascular effects or that might affect absorption, metabolism and excretion of nimodipine. Substantial procedures documented will include surgeries, ventricular catheter, lumbar puncture or drain, tracheostomy, gastrostomy and neurointerventional procedures for angiographic vasospasm or DCI.

### Endpoint Adjudication, Data Management and Data Monitoring Committees

An independent, blinded EAC will review all clinically significant episodes of hypotension, regardless of suspected relatedness to IP. The committee consists of 4 neurocritical care physicians who are independent from the Sponsor. They will be blinded to the subject’s study group. The EAC will decide if the hypotension is related to IP or not (yes or no). Two committee members will review each case and if they disagree, a third member will make the deciding assessment.

Independent contract research organizations (WuXi Clinical, Austin, TX, USA; Anju, Phoenix, AZ, USA; Suvoda, Conshohocken, PA, USA) are responsible for trial management. They conduct site recruitment and set up tasks, provide interactive response technology, site monitoring, manage electronic data capture and security and drug safety.

A Data Monitoring Committee (DMC) will review the safety of GTX-104 and oral nimodipine when 25 and 50 subjects have completed the treatment period. They are 3 individuals (neurocritical care and neurosurgery) who are independent from the Sponsor. A decision about subsequent reviews will be made after 50 patients are reviewed.

Prior to the start of the trial, the EAC and DMC had charters ratified that define the committee membership, roles, and responsibilities, meeting organization, format and materials to be provided and reviewed and meeting reports and recommendations.

### Statistics

The planned sample size is 100 subjects, with 50 in each arm. This sample size is not intended to provide adequate statistical power for specific comparisons or hypothesis tests of GTX-104 and oral nimodipine for any of the study endpoints. With a sample size of 50, an event with an incidence of 15% will have an exact (Clopper-Pearson) 95% CI width of approximately 0.22, with a lower limit of 0.065, and an upper limit of 0.279. The 95% CI width at 50% incidence will be approximately 0.29. The primary and all secondary endpoints will be presented based on a safety analysis set defined as all enrolled subjects according to their randomized group who receive at least 1 dose of IP after randomization. For the primary endpoint, the proportion of subjects with clinically significant hypotension determined by the EAC to have a reasonable possibility of relatedness to IP will be presented by the as randomized study group (ie. safety analysis set). No missing data imputation will be applied. The 2-sided 95% CI for the proportion will be obtained using the exact (Clopper-Pearson) method. The difference in the proportions between the two treatments and the exact 95% CI will also be presented.

Since the protocol allows GTX-104 patients to switch from IV to standard of care oral nimodipine, it is planned to analyze some subjects randomized to GTX-104 based on whether they are receiving IP or standard of care oral nimodipine prior to the occurrence of the endpoint (modified safety analysis set). This is planned only for the secondary endpoints of duration and number of clinically significant hypotension events and the incidence and severity of adverse events. The other secondary endpoints and the clinical and health economic outcomes will only be summarized by the randomized treatment group since they are less likely to be affected by the change in formulation of nimodipine being administered.

If the Day 90 quality of life and modified Rankin scale are missing, the Day 30 observation, if available, will be carried forward.

Duration of treatment with IP, total mass of standard of care oral nimodipine prior to randomization, total mass of IP during the treatment period and total mass of oral nimodipine after the switch from IV to oral during the treatment period, will be summarized. For each of the IP, the percent of the prescribed dose administered during the treatment period and a relative dose intensity will be calculated. The relative dose intensity will be defined as 100 x total mass of IP administered/total mass of IP that should have been administered.

### Protocol Amendments

This protocol incorporates the 5th amendment. **Tables 2 to 6** summarize the changes for each amendment.

**Table 2:** Changes Incorporated into Protocol Amendment 1.0.

| Section of Protocol | Original Protocol | Change to Protocol and Rationale |
| --- | --- | --- |
| **Objectives and Endpoints.**<br>**Primary Endpoint** | Incidence (% or proportion of subjects) with grade $\geq 2$ arterial hypotension possibly, probably, or definitely related to GTX-104/oral nimodipine. | Incidence (% or proportion of subjects) with at least 1 episode of grade $\geq 2$ arterial hypotension with a reasonable possibility that GTX-104/oral nimodipine caused the event, according to the blinded endpoint adjudication committee (EAC).<br>The EAC assessment of hypotension will be the primary endpoint since the EAC will be blinded to the treatment assignment. This will help mitigate bias. The question/criteria to determine relationship to Investigational Product was clarified based on FDA Guidance “Safety Reporting Requirements for INDs and BA/BE Studies”, December 2012. |
| **Objectives and Endpoints.**<br>**Secondary Endpoints** | Hemodynamic parameters (severity, duration and causality): systolic, diastolic and mean blood pressure and heart rate.<br>Grade $\geq 2$ arterial hypotension (causality, duration, incidence [number of episodes, number (%) of subjects]).<br>Adverse Events.<br>Use of concomitant medications, rescue therapy and endovascular interventions for angiographic vasospasm and delayed cerebral ischemia.<br>Other vital sign assessments (body temperature and respiratory rate). | These were reordered, simplified and clarified to:<br>Duration and total number of episodes of grade $\geq 2$ arterial hypotension.<br>Incidence and severity of Adverse Events (based on the National Cancer Institute Common Terminology Criteria for Adverse Events (NCI-CTCAE, Version 5.0).<br>Incidence of delayed cerebral ischemia and rescue therapy.<br>Suicidal ideation using the C-SSRS of $\geq 4$.<br>The rationale is the hemodynamic parameters, other vital sign assessments and laboratory assessments are vague and not defined. These parameters/assessments are combined into the new secondary endpoint: Incidence and severity of Adverse Events based on the National Cancer Institute Common Terminology Criteria for Adverse Events (NCI-CTCAE, Version 5.0). This amendment clarifies that clinically significant changes to these parameters or assessments will be reported as Adverse Events. |
|  | Suicidal ideation using the Columbia-Suicide Severity Rating Scale (C-SSRS).<br>Laboratory assessments (serum chemistry, hematology, urine analysis). | The C-SSRS score will be dichotomized into $<4$ or $\geq 4$ for each time it is assessed and then summarized by study arm because this cut-point in the scale defines those subjects who should be referred for immediate medical/psychiatric assessment. |
| **Exclusion Criteria** | Has received more than 3 doses of oral nimodipine (as a solution or capsules) as part of standard of care (SOC) for the ruptured aneurysm or within the previous 10 days before consideration for study entry and randomization into this clinical trial.<br>Is receiving cimetidine, or a strong inhibitor of CYP3A4, or a strong inducer of CYP3A4<br>Note: Use of cimetidine, or a strong inhibitor of CYP3A4 or a strong inducer of CYP3A4 prior to subarachnoid hemorrhage (SAH) and not within 5 times the drug's half-life prior to randomization is allowed. | Has received more than 5 doses of oral nimodipine (as a solution or capsules) as part of SOC for the ruptured aneurysm prior to randomization.<br>The rationale is that allowing only 3 doses of oral nimodipine gives 16 hours of time. Since most sites start oral nimodipine as soon as aneurysmal SAH is diagnosed, this time would make it almost impossible to randomize any subjects into the study.<br>Is receiving strong inhibitors of CYP3A4 such as some macrolide antibiotics (e.g., clarithromycin, telithromycin), some anti-HIV protease inhibitors (e.g., delaviridine, indinavir, nelfinavir, ritonavir, saquinavir), some azole antimycotics (e.g., ketoconazole, itraconazole, voriconazole) and some antidepressants (e.g., nefazadone).<br>The rationale is exclusion criteria related to CYP3A4 inhibitors and inducers were changed to be consistent with the FDA label. |
| **Hypotension and Dose Reduction** | Clarified and added text for hypotension with regard to dose reduction of investigational product (IP). | Revised Hypotension and Dose Reduction section to write for this study, 3 grades of hypotension based on blood pressure (BP) and medical intervention have been defined as follows:<br>Decrease in systolic BP $> 20$ mm Hg or diastolic BP $> 10$ mm Hg, or a systolic BP $\leq 100$ mm Hg, lasting for at least 5 minutes:<br>• Grade 1: not requiring any medical treatment |
|  |  | (pharmacotherapy or other intervention).<br>• Grade 2: requiring medical treatment without vasopressors, such as fluids, plasma or albumin perfusion, postural changes, dose reduction of IP or interruption of antihypertensive medications.<br>• Grade 3: requiring prescription of vasopressors or increase in dose (by more than 10%) of vasopressor or addition of a new vasopressor.<br>Doses of IP can be modified, when necessary, based on individual subject tolerance to treatment. Every effort should be made to avoid dose reduction of IP. If required, dose reduction/discontinuation should proceed as follows:<br>For subjects experiencing Grade 1 hypotension, no alteration in dose is recommended.<br>For subjects experiencing Grade 2 hypotension:<br>Dose regimens should be reduced as follows: For GTX-104: 30 minute intravenous (IV) bolus reduced to 2 mg every 4 hours (the background continuous IV infusion remains 0.15 mg/hour). For oral nimodipine: reduced to one 30 mg capsule every 4 hours. When at least 8 hours have elapsed since the hypotension resolves, the dose of IP may be increased to the full recommended dose, based on a medically-qualified investigator's decision.<br>For subjects experiencing Grade 3 hypotension: Treatment should be temporarily stopped. When at least 8 hours have elapsed since the hypotension resolves and there have not been escalating vasopressor requirements, dosing may be resumed at 50% of the initial dose, based on a medically-qualified investigator's decision: For GTX-104: 30 minute IV bolus of 2 mg every 4 hours (the background continuous IV infusion remains 0.15 mg/hour). For oral nimodipine: one 30 mg capsule every 4 hours. After at least 8 hours of tolerating the above regimen, the dosing of |
|  |  | IP may be increased to 100% of the initial dose, based on a medically qualified investigator's decision. For GTX-104: continuous IV infusion of 0.15 mg/hour and a 30 minute IV bolus of 4 mg every 4 hours. For oral nimodipine: two 30 mg capsules every 4 hours.<br>Note 1 was deleted. It wrote that by definition, subjects with grade 3 hypotension receive vasopressor(s). Therefore, resolution to grade 1, baseline, or to within normal range is considered after 12 hours of stable or reduced dose of vasopressors.<br>The rationale is that definitions of hypotension were added to this section and the instructions for dose reduction/stopping were clarified. The hypotension definitions are called grades 1 to 3 but these are not the grades used for assessing severity of adverse events. |
| **Concomitant Medications and Procedures** | Updated requirements regarding concomitant medications and interventions to harmonize with FDA label. | As written above, concomitant administration of IP (either GTX-104 or oral nimodipine) and strong CYP3A4 inhibitors is contraindicated. Strong inhibitors of CYP3A4 include some macrolide antibiotics (e.g., clarithromycin, telithromycin), some anti-HIV protease inhibitors (e.g., delaviridine, indinavir, nelfinavir, ritonavir, saquinavir), some azole antimycotics (e.g., ketoconazole, itraconazole, voriconazole) and some antidepressants (e.g., nefazadone). After intake of grapefruit juice and nimodipine, the blood pressure lowering effect may last for at least 4 days after the last ingestion of grapefruit juice. Ingestion of grapefruit / grapefruit juice is therefore not recommended while taking nimodipine.<br>If nimodipine is concomitantly administered with moderate and weak inhibitors of CYP3A4, BP should be monitored and if hypotension occurs, a reduction of the IP dose may be necessary.<br>Concomitant administration of other calcium channel blockers should be avoided, titrated down, or limited to the |
|  |  | first few hours or days of nimodipine therapy. Concomitant administration of other anti-hypertensive agents may require dose adjustment of such agents. Concomitant interventions are common in subjects with SAH, including but not limited to intensive care or hospital (re)admission, ventricular drainage, lumbar puncture, mechanical ventilation, use of nasogastric or gastric tubes and tracheostomy. Use of concomitant interventions will be captured in the electronic case report form.<br>The rationale is the contraindications for the strong CYP3A4 inhibitors and the intake of grapefruit juice are now consistent with the approved product information. |
| **Data Monitoring Committee** | Added a separate Data Monitoring Committee (DMC). | An independent, blinded Endpoint Adjudication Committee (EAC) will review all episodes of hypotension, regardless of suspected relatedness to IP. The DMC will review the safety of GTX-104 and oral nimodipine according to a pre-specified schedule, either on a calendar basis or based on the number of subjects treated.<br>Members of the EAC and DMC will each consist of 3 to 5 independent experts with relevant medical and scientific experience. Prior to the start of the trial, the EAC and DMC will have charters ratified. |

**Table 3:** Changes Incorporated into Protocol Amendment 2.

| **Section of Protocol** | **Existing Protocol** | **Change to Protocol and Rationale** |
| --- | --- | --- |
| **Primary and Secondary endpoints** | Incidence (% or proportion of subjects) with at least 1 episode of grade $\geq 2$ arterial hypotension with a reasonable possibility that GTX-104/oral nimodipine caused the event, according to the blinded | Incidence (% or proportion of subjects) with at least one episode of clinically significant hypotension with a reasonable possibility that GTX-104/oral nimodipine caused the event, according to the blinded EAC.<br>The grades and definitions of hypotension were changed. |
|  | Endpoint Adjudication Committee (EAC). | The rationale is that the grading system was complicated and not clinically relevant. Essentially the only important hypotension is that which the investigator does something about, which is now defined as clinically significant hypotension. The primary endpoint has been updated to incidence (% or proportion) of subjects with at least 1 episode of clinically significant hypotension. |
| **Clinical and Health Economic Outcomes** |  | Added modified Rankin Scale (mRS) to be assessed at Day 30 and Day 90.<br>The rationale is that the mRS is the most common functional outcome scale used to assess outcomes after stroke, including after aneurysmal subarachnoid hemorrhage (aSAH). It has been added as a general assessment at up to 3 months after the aSAH of the effects of any adverse events that occur. |
| **Hypotension and Dose Reduction** | This is described in Table 2 under the **Hypotension and Dose Reduction** section 3<sup>rd</sup> column (**Change to Protocol and Rationale**) | For this study, hypotension is defined as follows:<br>Decrease in systolic blood pressure (BP) > 20 mm Hg or diastolic BP > 10 mm Hg, or a systolic BP ≤ 100 mm Hg, confirmed by 2 consecutive readings within 5 minutes.<br>The rationale is to clarify how BP readings should be confirmed and for consistency with other sections of the protocol.<br>Two categories of hypotension have been defined as follows:<br>1. Not clinically significant: not requiring any medical treatment (pharmacotherapy or other intervention).<br>2. Clinically significant: requiring medical treatment, including but not limited to intravenous fluids, postural changes, dose reduction of IP, interruption of antihypertensive medications, prescription of vasopressors, increasing dose of a vasopressor or addition of a new vasopressor.<br>Hypotension definitions were collapsed to 2 categories that are consistent with the primary endpoint: not clinically significant (previously grade 1, defined as not treated) and clinically significant (previously grade 2 or 3, now collapsed to one |
|  |  | category since they are both hypotension events that are treated). The terminology was changed from “grade” to "not clinically significant/ clinically significant" to avoid confusion with severity grading used for adverse events.<br>The dose reduction guidelines have been modified to align with the two categories of hypotension and to delete the option to stop/restart the IP. Stopping IP is always an option in any clinical trial and doesn’t need to be specified. The section now reads:<br>Since nimodipine is administered as standard of care (SOC) to patients with aSAH, every effort should be made to give the entire prescribed dose of investigational product (IP) and to avoid dose reductions or interruptions of IP.<br>If deemed necessary based on investigator decision, dose reduction of IP should proceed according to SOC. The recommended dose reductions may be: For GTX-104: 30 minute IV bolus of 2 mg every 4 hours. The background of continuous infusion remains 0.15 mg/hr. For oral nimodipine: One 30 mg capsule every 4 hours.<br>The rationale to recommend these dose reductions was based on the fact that this is basically SOC at many centers even though this is not recommended in the prescribing information for oral nimodipine capsules. |
| **Suicidal Ideation and Behavior Risk Monitoring** | Suicidal Ideation and Behavior Risk Monitoring | The Columbia-Suicide Severity Rating Scale (C-SSRS) Initial or Baseline Form will also be completed at screening (if subject is capable) or the first time the assessment is completed.<br>The rationale is that this is consistent with how the C-SSRS is used in clinical practice. The baseline form should be completed the first time the scale is performed so that the lifetime scale is obtained.<br>The protocol also added that if a subject has a score $\geq 4$ on the C-SSRS or evidence of suicidal behavior is noted, the subject should be referred to a mental health professional for evaluation and |
|  |  | potential treatment.<br>The rationale is standard, proper medical care and patient safety for any subject with active suicidal ideation. If a subject has a score $\geq 4$ on the C-SSRS or evidence of suicidal behavior is noted, the subject should be referred to a mental health professional for evaluation and potential treatment. |

**Table 4:** Changes Incorporated into Protocol Amendment 3.

|  |  |  |
| --- | --- | --- |
| **Columbia-Suicide Severity Rating Scale (C-SSRS)** | If a subject has a score $\geq 4$ on the C-SSRS, or evidence of suicidal behavior is noted, the subject should be referred to a mental health professional for evaluation and potential treatment. | If a subject has a score $\geq 4$ or has a score that indicates active suicidal ideation or behavior, on the C-SSRS, the subject's mental health practitioner will be contacted immediately (if applicable) or the subject will be directed to the emergency department. Should a subject exhibit suicidal ideation or behavior while in hospital or have scores on the C-SSRS that reflect such thoughts, appropriate evaluation and treatment by a mental health professional while hospitalized should occur.<br>The rationale is to bring the management of patients with a score $\geq 4$ into agreement with best medical practice. |
| **Blood Pressure (BP) and Heart Rate** | Not mentioned. | Added to the protocol was that BP will be captured in the electronic case report forms when subjects demonstrate signs concerning for hypotension (e.g. dizziness, lightheadedness, excess somnolence).<br>The rationale was that the FDA was concerned we would miss symptomatic hypotension episodes. |

**Table 5:**
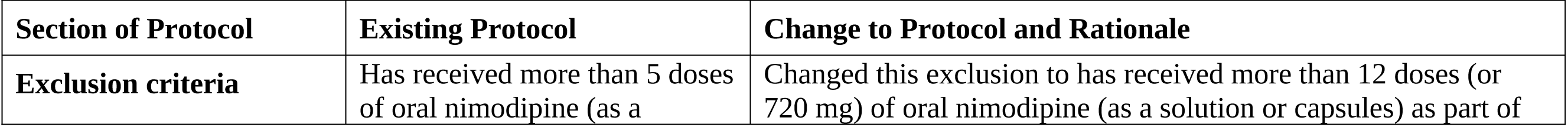
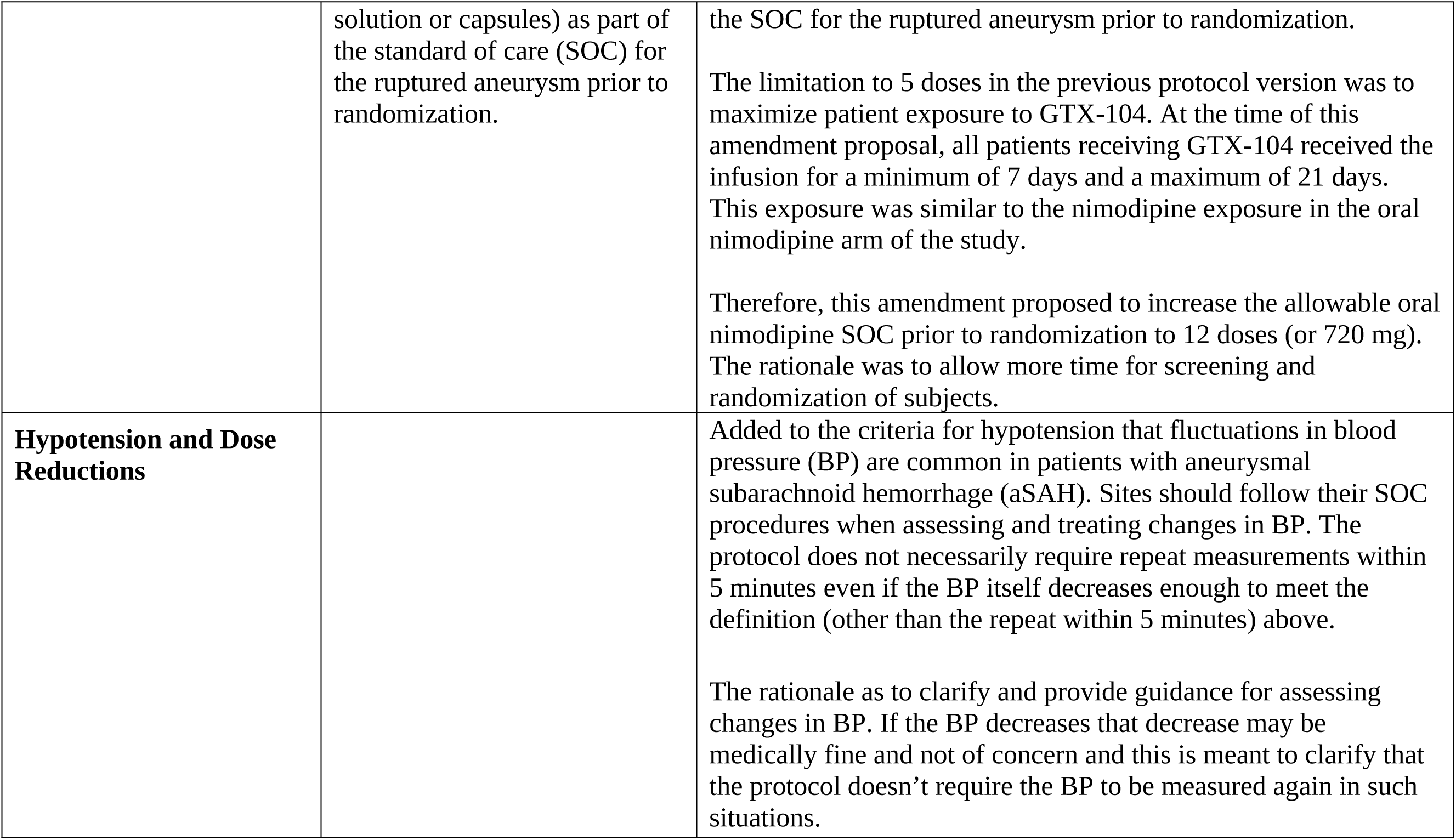
Changes Incorporated into Protocol Amendment 4.

**Table 6:** Changes Incorporated into Protocol Amendment 5.

| Section | Description of Change | Summary of Changes and Rationale |
| --- | --- | --- |
| **Hypotension and Dose Reduction** | Changes to blood pressure (BP) evaluation and dose reductions. | A statement was added to clarify evaluation of not clinically significant hypotension events: Fluctuations in BP are common in patients with aneurysmal subarachnoid hemorrhage (aSAH). Sites should follow their standard of care (SOC) when assessing and |
|  |  | treating changes in BP. If BP drops and meets the definition of hypotension above, is repeated within 5 minutes and is not treated, these events should be reported as not clinically significant. In the event that the BP measurement is not repeated within 5 minutes yet medical treatment is rendered, the event should be considered clinically significant.<br>The rationale is that since the SOC at most sites is to repeat measurement of BP every 15 minutes in the first hours to days after aSAH, text has been added to address the situation in which there has not been two measurements of BP within 5 minutes. In this situation if the first reading results in treatment of hypotension, the sites should record the event as clinically significant and the event should be adjudicated by the Endpoint Adjudication Committee.<br>The dose reduction recommendations also were revised again. The protocol now states that if deemed necessary based on investigator decision, dose reduction should proceed according to the US label (Nimodipine Package Insert). The recommended dose reductions may be: For GTX-104: 30 minute intravenous (IV) bolus of 2 mg every 4 hours. The background continuous IV infusion remains 0.15 mg/hour. For oral nimodipine: One 30 mg capsule every 4 hours. Information about dose reduction and discontinuation will be captured in the electronic case report forms.<br>The rationale to remove the recommended dose reductions to dosing every 2 hours (30 minute IV bolus of 2 mg every 2 hours or one 30 mg capsule every 2 hours) was based on FDA recommendations that the dose regimen follow the US label for oral nimodipine which does not mention dosing every 2 hours. |

## DISCUSSION

Intravenous nimodipine could have some advantages over oral nimodipine capsules. However, the safety and efficacy of nimodipine for aSAH relies heavily on one randomized clinical trial that used oral nimodipine tablets, 60 mg every 4 hours.^26^ Gelatin capsules are used in the US and tablets in Canada. The dosages of each are the same but the bioavailability and peak plasma concentrations are higher with the gelatin capsules compared to the tablets.^37^ The most important side effect of nimodipine is hypotension, which is closely associated with peak plasma concentrations greater than 30-40 ng/mL.^12,20,38^ In healthy volunteers, this can be relatively easily controlled by adjusting the dose. However, plasma concentrations of nimodipine fluctuate more widely in patients with aSAH because of the variability in absorption and metabolism that occurs in these patients. The plasma concentration of oral nimodipine depends on the dose, dosage form, stomach contents, CYP3A4 activity in the small intestine wall and liver (first pass effects), liver function, the patient’s cardiovascular condition and what BP parameters are targeted for the patient. More complexity is added because many of these things change over time in aSAH patients. Intravenous nimodipine, however, is not affected by as many factors.

Food and first pass effects are probably the most important source of nimodipine plasma concentration variability. Bioavailability of oral nimodipine ranges from 3 to 28% in patients with aSAH and from 5-13% in healthy volunteers.^37–41^ This suggests that by avoiding this variability, hypotension could be less common with IV nimodipine. Studies of GTX-104 in healthy human volunteers found less variability in PK with GTX-104 compared to oral nimodipine (unpublished data, Acasti Pharma). Furthermore, gastrointestinal side effects like diarrhea are less common or don’t occur with IV nimodipine.^5^

Additional rationale for use of IV nimodipine are ease of use since it does not require patients to swallow the capsules or for the contents of capsules to be extracted for injection down gastric tubes. This could lead to improved compliance. There is no risk of inadvertent IV injection that is a black box warning with the capsules. GTX-104 also does not contain any preservatives or vessel-irritating components like ethanol so it can be injected through a peripheral venous catheter if desired. This also makes it theoretically better for intrathecal/intraventricular use than nicardipine which contains preservatives and for intraarterial use than the currently available IV nimodipine solution.

Hypotension is the primary endpoint of this study and the only important side effect of nimodipine. Hypotension is believed to be harmful to patients with aSAH especially from 3 to 14 days after the hemorrhage.^2,42^ When it occurs, the dosage of nimodipine often is reduced or nimodipine is stopped altogether. This is problematic because administration of the full recommended nimodipine dose has been associated with better outcome in some but not all reports.^7,43,44^ Multivariate analysis of 220 aSAH patients who were given oral nimodipine found only 96 (44%) got the full dosage for the first 14 days post aSAH. The dose was reduced to 50% in 63 (29%) patients and discontinued altogether in 61 (28%). Multivariate analysis found unfavorable outcome was associated with increased age, worse Hunt Hess grade and receiving less nimodipine.^7^ Another study of 309 aSAH patients found oral nimodipine dose was reduced or halted for at least 24 hours between days 5 and 10 post-SAH in 108 (53%) patients.^44^ Patients who had dose reductions or stoppages were more likely to be poor grade and have higher Fisher scores on admission computed tomography. In multivariate analysis the only factor associated with DCI was reduced nimodipine dose. Factors associated with poor outcome at 3 months follow-up were age, poor neurological grade and reduced nimodipine dose. Other studies have associated DCI with reduced nimodipine dose.^4,19^

Most published data on IV nimodipine is from retrospective case series and prospective phase 2 studies.^45–50^ There are 2 prospective randomized clinical trials comparing IV and oral nimodipine with placebo.^51,52^ Both prescribed IV nimodipine for 7 to 14 days followed by oral nimodipine. Jan, et al., randomized aSAH patients within 24 hours of neurological deterioration from vasospasm so this was a treatment not prophylaxis study.^51^ Nimodipine was associated with better clinical outcome, although the analysis was after excluding 61 of 188 randomized patients. Most of the patients in Ohman, et al., were randomized and started on nimodipine within 72 hours of aSAH. There was no benefit of nimodipine on outcome at 3 months.^52^ Thus, no conclusions can be drawn about the efficacy of continuous infusions of IV nimodipine.

Two randomized clinical trials compared oral and IV nimodipine and found no difference in clinical outcome but the numbers of patients are too small to show noninferiority or differences in efficacy.^53,54^ No studies of IV nimodipine used the dose regimen that is given in this study. This dose regimen was chosen because it replicates the pharmacokinetic profile of oral nimodipine. Whether this profile is important for the efficacy of nimodipine is unknown but this regimen fulfills the pharmacokinetic bridging usually required for this type of study.

## Data Availability

This manuscript does not include any data.

## Author Contributions

The protocol was written, revised and amended with input from A Choi, SH-Y Chou, AF Ducruet, WT Kimberly, RL Macdonald and AA Rabinstein. RL Macdonald wrote the manuscript.

